# Evaluation of the Efficacy and Adverse Effects of Isotretinoin in the Treatment of Acne Vulgaris, with a Focus on Psychological Impact in Low-Income Countries: A Cross-Sectional Study from Syria

**DOI:** 10.1101/2025.09.03.25335066

**Authors:** Aya Barakat, Raghad Merai, Alaa Alhaboul, Musab Murad, Sophie Barguil

**Affiliations:** Faculty of Pharmacy, Damascus University, Syria; Department of Toxicology and Pharmacology, Faculty of Pharmacy, Damascus University, Syria

## Abstract

**Background and Aim:** This study aims to evaluate the effectiveness of isotretinoin in the treatment of acne, while also monitoring its associated physical and psychological adverse effects.

**Materials and Methods:** A descriptive cross-sectional study was conducted using an online questionnaire distributed to individuals who are currently experiencing acne or have experienced it in the past. The questionnaire included items regarding acne severity, type and duration of treatment, adherence, side effects, and overall satisfaction. Data were statistically analyzed.

**Results:** A total of 377 participants, aged between 21 and 25 years, were included In the study. A significant Improvement was reported by 74.5% of users following isotretinoin treatment, while 4.8% noticed no improvement.Side effects were experienced by 91.2% of the participants.

**Physical side effects:** The most commonly reported side effects were dryness— particularly of the lips, skin and eyes (82.8%, 75.9%, and 45.4%, respectively).

**Psychological side effects:** 46.7% of participants reported symptoms such as anxiety, mood changes, depression, or even suicidal Ideation. Despite the side effects, 86.2% were satisfied with the treatment’s efficacy, while 19.9% discontinued the medication due to adverse effects.

**Conclusions:** Isotretinoin is widely regarded as an effective treatment for moderate to severe cane. However, its use requires strict medical supervision before and during the treatment period due to Its wide range of potential physical and psychological adverse effects.

The findings underscore the critical importance of comprehensive awareness among patients, physicians, and pharmacists regarding the potential risks—particularly psychological ones—that may escalate to suicidal tendencies in vulnerable individuals.

This highlights the necessity for integrated management approaches that include regular medical and psychological monitoring.

Given the persistent neglect of mental health In many communities, these results are concerning and call for increased public awareness and a shared responsibility among healthcare providers and patients.

As for physical symptoms such as various forms of dryness, they can be detected early and managed medically. In some cases, dose adjustments or discontinuation may be necessary to prevent serious complications.

## Introduction

The skin is composed of three primary layers: the epidermis, dermis, and subcutaneous tissue, each of which plays a vital role in protecting the body and maintaining homeostasis [1].

Among the most common disorders affecting this structure is acne vulgaris, a chronic inflammatory condition of the pilosebaceous units. It is particularly prevalent among adolescents and manifests as various types of lesions, including comedones, papules, pustules, nodules, and in some cases, painful inflammatory cysts. These lesions commonly appear in areas rich in sebaceous glands, such as the face, chest, back, and shoulders [2].

According to the American Academy of Dermatology (AAD), approximately 85% of individuals aged 12 to 24 experience some form of acne, although cases have also been documented in children and older adults [3].

Despite that the acne is not classified as a life-threatening condition, its psychological and social impact is substantial, often affecting self-esteem, social interaction, and overall quality of life. As a result, growing attention has been paid to identifying treatments that are not only effective but also safe and well-tolerated [2].

Isotretinoin, a synthetic vitamin A derivative, remains one of the most effective therapies for moderate to severe acne, however, its use is frequently associated with a range of adverse effects including mucocutaneous symptoms, systemic side effects [4].

Recently a growing concern regarding potential psychiatric manifestations such as depression, anxiety, and suicidal ideation. Numerous studies have addressed the efficacy and safety profile of isotretinoin from various clinical perspectives.

This study contributes to the existing body of research by placing particular emphasis on the psychological and physical adverse effects observed in a Syrian population. Through its focus on patient-reported outcomes, adherence patterns, and the influence of treatment source and education, it seeks to highlight aspects that may support more holistic and context-specific approaches to acne management.

## Materials and Methods

### I. Study Design and Population

This cross-sectional descriptive study was conducted between November 2024 and April 2025 and aimed to assess the physical and psychological adverse effects of isotretinoin among individuals who had previously used or were currently using the medication to treat acne vulgaris.

The study included a purposive sample of 377 participants from across Syria, selected to represent a diverse population in terms of age, gender, and social background.

### II. Data Collection Tool

A structured electronic questionnaire was developed specifically for this study, building upon validated tools used in prior Syrian and international research [5,6].

The questionnaire was reviewed by a panel of academic experts in toxicology and pharmacology to ensure content validity and relevance to the local context.

It comprised both closed- and open-ended questions covering: Demographics and clinical history, Duration and severity of acne, Duration of isotretinoin use,Degree of adherence, Physical side effects (e.g., mucocutaneous symptoms, headaches, visual disturbances), Psychological symptoms (e.g., mood swings, depression, suicidal ideation), Source of medical supervision,Overall satisfaction with treatment.

The questionnaire was distributed via targeted digital outreach through student medical networks, dermatology-related forums, and academic social media groups affiliated with Damascus University. This approach ensured access to individuals likely to have relevant experience with isotretinoin treatment while maintaining a degree of sampling specificity.

### III. Ethical Considerations

The study protocol was reviewed and approved by the Ethics Committee of the Faculty of Pharmacy, University of Damascus. All participants provided informed digital consent before completing the questionnaire. Participation was entirely voluntary and anonymous. No personal identifiers were collected, and strict confidentiality of all responses was maintained throughout the study, in accordance with the ethical principles outlined in the Declaration of Helsinki.

### IV. Statistical Analysis

All data were analyzed using the Statistical Package for the Social Sciences (SPSS).

Descriptive statistics were used to summarize participant characteristics, treatment patterns, and reported side effects. The Chi-square test was applied to evaluate associations between categorical variables. A p-value of less than 0.05 was considered statistically significant.

## Results

A total of 377 participants completed the questionnaire. The majority were female (90.7%) and aged between 21 and 25 years (65%). The acne severity among participants was most commonly reported as moderate (58.6%).

Regarding isotretinoin use, 89.1% of participants had taken the medication, with 81.7% doing so under dermatological supervision. However, 70.7% did not maintain regular follow-up with their physician. The duration of treatment varied, with 35.3% using the drug for four to six months.

### Treatment Outcomes and Adverse Effects

A significant improvement in acne symptoms was reported by 74.5% of participants, while 4.8% experienced no improvement, and 1.3% reported worsening of their condition. Notably, 91.2% of users experienced at least one side effect.

The most commonly reported physical adverse effects were:

– Dry lips: 82.8%
– Dry skin: 75.9%
– Dry eyes: 45.4%

Psychological symptoms were also prominent, with 46.7% of participants reporting experiences such as anxiety, mood swings, depression, or suicidal thoughts.

Despite these adverse events, 86.2% of users expressed satisfaction with the treatment, while 19.9% discontinued the medication due to side effects.

### Key Statistical Associations

- Source of treatment vs. level of adherence: χ ^2^ = 36.086, p = 0.00012 (table 1)
- Receiving adequate information vs. discontinuation of treatment: χ ^2^ = 6.657, p = 0.010 (table 2)
- Adherence level vs. presence of side effects: χ ^2^ = 0.205, p = 0.977(table 3)
- Age group vs. therapeutic efficacy: χ ^2^ = 5.602, p = 0.779 (table 4)
- Presence of side effects vs. discontinuation of treatment: χ ^2^ = 0.236, p = 0.627 (table 5)
- Acne severity vs. therapeutic efficacy: χ ^2^ = 9.699, p = 0.138 (table 6)
- Duration of treatment vs. occurrence of side effects: χ ^2^ = 6.711, p = 0.152 (table 7)
- Age group vs. psychological manifestations: χ ^2^ = 3.603, p = 0.308(table 8)

## Discussion

Although numerous studies have explored this field in terms of pharmacological efficacy and side effects, this study sheds light—in addition to the above—on the psychological effects.

While these effects are no less important than physical ones, they are often neglected for one reason or another, especially in developing societies.

The survey shows that 89.1% of the 377 participants used isotretinoin for acne treatment, a significant and notable percentage indicating the drug’s widespread use. The majority had moderate-severity acne lesions, but 74.5% reported improvement.

However, 18.3% did not follow any medical guidance and were not under medical supervision. In developing countries, some prescription-only drugs are dispensed over-the-counter (OTC).

Patients, believing they were treating superficial skin lesions, were not fully aware of the drug’s harmful effects.

This is evidenced by the 1.3% whose condition worsened instead of improving—all of whom had taken the drug without a prescription, as confirmed by questioning in this study.

Compared to Western studies, the rates of mucocutaneous side effects in the studied sample were higher. For example, 82.8% experienced lip dryness, compared to 60–70% in a 2014 British clinical study (though the difference was not statistically significant) [7].

Complaints of dry skin and eyes were also notably more frequent. This discrepancy may be attributed to environmental factors in Syria, genetic predispositions in the population, or differences in daily skincare and moisturizing habits.

However, the most striking observation in this study’s results pertains to the psychological aspect: 46.7% of participants reported symptoms such as mood changes or depression—a significantly higher percentage than the 20–30% typically recorded in European and American studies.

For instance, a 2021 systematic review published in Dermatologic Therapy [8] reported lower rates. However, this aligns with a 2019 Syrian study (55.6% for mood changes), showing no significant statistical difference [5].

This percentage is concerning, especially given the widespread OTC use of isotretinoin in our society, often taken randomly without precise medical supervision, reflecting a lack of awareness of the drug’s potential psychological risks.

Moreover, ignoring or excluding psychological support during treatment—despite studies proving the drug’s potential to induce depression—increases the likelihood of worsening psychological effects in users. This lack of awareness and follow-up poses a real challenge in mitigating potential adverse effects, particularly in settings where integrated mental healthcare is absent from treatment plans.

The correlation between treatment source and adherence severity suggests that patients obtaining treatment from reliable sources (e.g., specialists or healthcare centers) show higher compliance. This is attributed to greater trust in the source, detailed explanations of dosage, side effects, and treatment duration, reinforcing the patient’s conviction in follow-up. Conversely, reliance on unverified sources (e.g., friends or social media) reduces adherence due to a lack of scientific guidance or skepticism about treatment efficacy.

## Limitations

While this study provides valuable insights into the physical and psychological adverse effects of isotretinoin among Syrian patients, several limitations must be acknowledged:

– Sample Demographics: The sample was predominantly female and composed mainly of young adults (aged 21–25), which, while representing a high-risk group for acne, may limit the generalizability of the findings to broader age and gender groups.
– Self-Reported Data: All responses were self-reported, making the study susceptible to recall bias or reporting bias. There was no objective clinical verification of acne severity or psychological manifestations, which could have strengthened the findings.
– Lack of Standardized Psychological Tools: The study did not employ validated psychological assessment instruments such as the PHQ-9 or Beck Depression Inventory. This limits the ability to compare psychological outcomes directly with international studies.
– Cross-Sectional Design: As with all cross-sectional studies, causal relationships cannot be established. The associations observed reflect correlations, not definitive cause-effect conclusions.
– Digital Distribution Bias: The questionnaire was distributed electronically, potentially excluding individuals with limited internet access or digital literacy, thereby influencing the representativeness of the sample.

## Conclusion

The study demonstrated that isotretinoin is an effective treatment for moderate to severe acne, but it is associated with physical and psychological side effects that require close monitoring. It also revealed that prior awareness of the risks enhances knowledge about the drug and its accompanying symptoms, underscoring the crucial role of pharmacists and medical guidance.

The findings highlight the need for stricter regulation of drug dispensing and the inclusion of psychological support in treatment plans, particularly in settings with insufficient awareness of psychological complications. This study sheds light on significant gaps in medical practice and calls for adopting a more comprehensive and safer therapeutic approach.

## Data Availability

All relevant data underlying the findings of this study are fully available within the manuscript and its Supporting Information files. This study was based on anonymous electronic survey responses, and all aggregated results are presented in the manuscript and included tables.

